# Reduction in antibiotic prescribing attainable with a gonococcal vaccine

**DOI:** 10.1101/2020.12.11.20247973

**Authors:** Stephen M. Kissler, Moriah Mitchell, Yonatan H. Grad

## Abstract

We estimated the fraction of antibiotic prescribing in the US attributable to gonorrhea. Gonorrhea contributes to an outsized proportion of antibiotic prescriptions in young adults, males, and in the southern and western US. A gonococcal vaccine could substantially reduce antibiotic prescribing in these populations.

## Introduction

Antibiotic resistance poses a growing threat to human health^1^. Antibiotic resistance is driven by the consumption of antibiotics, which exert evolutionary selective pressures on the intended target pathogen and on ‘bystander’ microorganisms^2^. Vaccination has been proposed as a means of reducing the incidence of illnesses that prompt antibiotic prescriptions, thereby reducing down-stream antibiotic resistance^3^.

Gonorrhea is the second most prevalent reportable illness in the United States, with nearly 600,000 cases reported in 2018^4^, and is a major contributor to morbidity worldwide^5^. The US treatment recommended by the Centers for Disease Control and Prevention (CDC) for gonorrhea is dual therapy with single dose oral azithromycin and intramuscular ceftriaxone^6^. Gonorrhea therefore contributes substantially to antibiotic prescribing.

There is currently no vaccine licensed for the prevention gonorrhea, though modest vaccine-induced protection has been demonstrated via cross-protection from the MeNZB *Neisseria meningitidis* vaccine and the investigation of the benefit of meningococcal group B vaccine (Bexsero) is ongoing^7,8^. While the impact an effective gonococcal vaccine could be profound in terms of reductions in gonorrhea-associated morbidity, it also has the potential benefit of reducing a large amount of antibiotic prescribing. Such a vaccine might reduce the threat of multi-drug-resistant gonorrhea – a top public health threat according to the CDC^1^ – as well as reduce the selective pressures exerted on other bystander organisms during the course of treatment for gonorrhea. However, the extent of impact of a gonococcal vaccine on antibiotic prescribing has not been quantified. Here, we integrate reported cases of gonorrhea in the United States with a large database capturing outpatient antibiotic prescriptions to estimate the amount of antibiotic prescribing attributable to gonorrhea and thus the reduction in antibiotic prescribing that would be possible with a gonococcal vaccine.

## Methods

### Study sample and population

We extracted annual cases of gonorrhea and chlamydia by state, sex, and age group (0–14, 15– 19, 20–24, 25–29, 30–34, 35–39, 40–44, 45–54, 55–64, 65+) in the United States from 2015-2018, the most recent years available, from the CDC’s *AtlasPlus* portal^9^. We used the population sizes for each demographic and geographic group, available from the same source, to calculate cases per 1,000 individuals (**Table 1**).

**Table 1.**
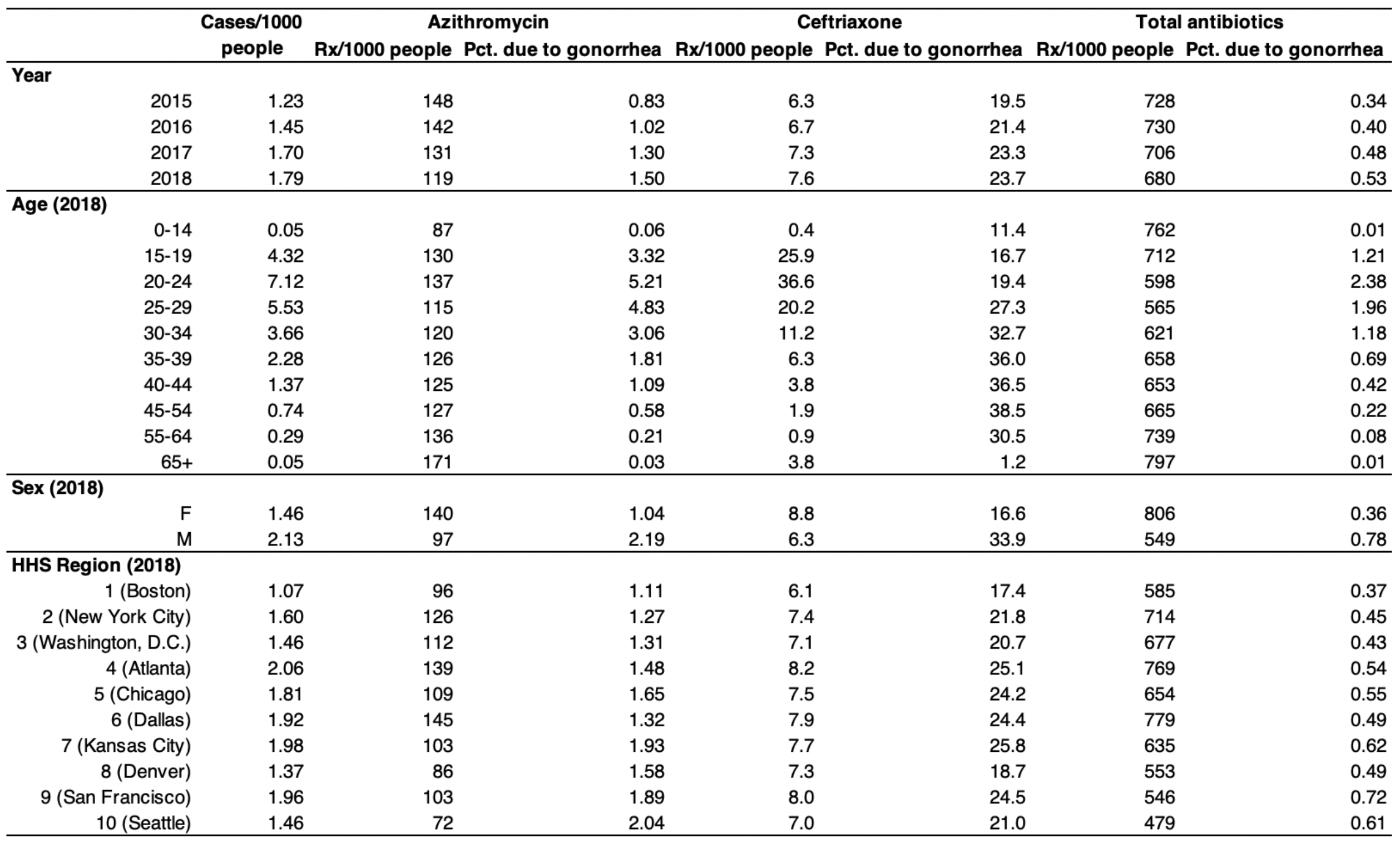
Gonorrhea cases and antibiotic prescribing rates per 1,000 people in the United States. Rx=prescriptions; pct=percent.

We extracted outpatient antibiotic prescriptions from the Truven MarketScan database^10^ over the same timeframe. This database captures all outpatient pharmacy claims from a convenience sample of 19.1–24.3 million individuals (5.9–7.6% of the US population), depending on the month. We linked each antibiotic prescription with the patient’s age, sex, state. Full extraction details are given in the **Supplement**.

Next, we estimated the number and proportion of azithromycin prescriptions, ceftriaxone prescriptions, and overall antibiotic prescriptions in the US that were attributable to gonorrhea in 2015-2018. The recommended course of azithromycin and ceftriaxone for treating gonorrhea is normally administered under the supervision of a provider upon diagnosis, so these prescriptions are not recorded in the MarketScan outpatient prescriptions data. We therefore added one dose of azithromycin and one dose of ceftriaxone for each reported gonorrhea infection to the outpatient antibiotic prescriptions data extracted from MarketScan. We also accounted for antibiotic prescriptions for chlamydia, the most common reportable illness in the US, by adding one dose of azithromycin to the MarketScan outpatient prescriptions data for each reported chlamydia case. This is a conservative choice, since chlamydia may be treated by a single supervised dose of azithromycin (recommended) or a 7-day course of doxycycline. By assuming that all cases of chlamydia prompt an azithromycin dose, this will lead to an over-estimate of the number of azithromycin prescriptions that are actually given, yielding more conservative estimates of the relative impact of a gonococcal vaccine on azithromycin prescribing.

For the years 2015–2018, we calculated the percent and total number of azithromycin prescriptions, ceftriaxone prescriptions, and total antibiotic prescriptions per 1,000 people that were attributable to gonorrhea. For 2018, when gonorrhea incidence was at its highest level in over a decade, we calculated the same statistics stratified by age, sex, and HHS region^11^.

## Results

There were 1.79 reported cases of gonorrhea per 1,000 individuals in the United States in 2018. These accounted for 1.5% of all azithromycin prescriptions, 23.7% of all ceftriaxone prescriptions, and 0.53% of all antibiotic prescriptions of any type given in 2018 (**Table 1**). The incidence of gonorrhea has been rising steadily, increasing by a factor of 1.5 between 2015 and 2018. Meanwhile, overall antibiotic prescribing rates have fallen, so that the proportion of antibiotic prescriptions due to gonorrhea has increased even faster than the increase in gonorrhea incidence alone would suggest. Reported gonorrhea cases are mainly concentrated in young adults. There were 7.12 cases of gonorrhea reported per 1,000 individuals between the ages of 20 and 24 in 2018, accounting for 2.38% of all antibiotic prescriptions given in that age group. Reported gonorrhea cases were less frequent in females than in males (1.46 *vs*. 2.13 cases per 1,000 individuals in 2018, respectively). Reported gonorrhea cases in 2018 were lowest in the northeast (1.07 cases per 1,000 people in HHS Region 1, Boston) and highest in the southeast (2.06 cases per 1,000 people in HHS Region 4, Atlanta). However, gonorrhea-related antibiotic prescriptions accounted for the greatest share of overall antibiotic prescriptions in the west (0.72% of all antibiotic prescriptions in HHS Region 9, San Francisco) due to a relatively high rate of gonorrhea infections and a relatively low rate of antibiotic prescribing for other conditions. A vaccine could prevent some fraction of the antibiotic prescriptions associated with gonorrhea cases. Such a vaccine would likely have the greatest impact on antibiotic prescribing in young adults, in males, and in the southern and western US.

## Discussion

We have estimated the share of antibiotic prescribing in the United States attributable to gonorrhea infections and have identified the sub-populations in which antibiotic prescribing would likely decline the most with an effective gonococcal vaccine. Gonorrhea incidence and associated gonorrhea-related antibiotic prescribing are especially variable between age groups, with the highest incidence and highest associated prescribing rates in young adults. Since overall antibiotic prescribing rates are lowest in this same age group, a gonococcal vaccine would yield an outsized benefit in this age group in terms of the fraction of antibiotic prescriptions averted. Reducing antibiotic prescribing in demographic groups that otherwise receive few antibiotics may be an especially effective strategy for combatting antibiotic resistance, since a given antibiotic course is expected to contribute more to resistance in an individual who receives few other prescriptions than in an individual who already receives many^12^. Taken together, these findings suggest that a gonococcal vaccine could lead to a substantial reduction in antibiotic prescribing, helping to combat the rise in antibiotic resistance.

Our findings align with others that have documented higher rates of gonorrhea in the southern United States, in males, and in young adults^4^. Similarly, elevated rates of antibiotic prescribing in the southern United States and in children and older adults have been documented^13^. Gonorrhea incidence has been rising rapidly over the past decade along with rates of antibiotic resistance in gonorrhea. We anticipate that this trend will continue, making gonorrhea account for an even larger share of antibiotic prescriptions and increasing the potential benefit of a gonococcal vaccine for reducing both gonorrhea-related morbidity and associated antibiotic prescribing.

The ultimate impact of a gonococcal vaccine will depend on the vaccine’s efficacy and its uptake in the population. Prioritizing vaccination in young adults would likely have the greatest benefits for both reductions in morbidity and reductions in antibiotic prescribing. Potential reductions in antibiotic prescribing should be considered when assessing the cost-effectiveness of a gonococcal vaccine. While we have focused on vaccination, other measures to reduce gonorrhea incidence, such as increased condom use, should also lead to a downstream reduction in antibiotic prescribing. We have focused on gonorrhea incidence in the United States, but we anticipate that a gonococcal vaccine would similarly lead to substantial reductions in antibiotic prescribing world-wide.

## Data Availability

Data are publicly available according to the citations within the manuscript.

## Supplement

### Extracting antibiotic claims from the MarketScan database

To extract antibiotic prescribing rates from the MarketScan database, we began by extracting the age, sex, state of enrollment, and member identification number for every person with a full month of insurance enrollment for each month between January 2015 and December 2018. **Supplemental Table 1** lists the number of members enrolled in each month and the proportion of the US population represented. For each member in each month, we then extracted all associated antibiotic claims. Antibiotics were identified by the National Drug Code (NDC) number using a lookup file containing the therapeutic class and drug name provided by Truven MarketScan. To estimate the annual *per capita* antibiotic prescribing rate, we divided the total number of antibiotics prescribed in a given year by the total number of member-months in that year (*i*.*e*., the sum of the number of members enrolled across all months in that year) and multiplied by 12. Multiplying this quantity by 1,000 yielded the number of antibiotic prescriptions per 1,000 people in that year. We performed these extractions for azithromycin prescriptions, ceftriaxone prescriptions, and all antibiotic prescriptions.

**Supplemental Table 1.** Number of Insurance members represented in the MarketScan database. The percent of the US population represented in the MarketScan data is calculated using the monthly resident population of the United States as reported by the United States Census Bureau (https://www.census.gov/data/tables/time-series/demo/popest/2010s-national-total.html)

| Year | Month | Members (millions) | Percent of US population |
| --- | --- | --- | --- |
| 2015 | 1 | 24.3 | 7.61 |
|  | 2 | 24.3 | 7.60 |
|  | 3 | 24.3 | 7.58 |
|  | 4 | 24.2 | 7.56 |
|  | 5 | 24.1 | 7.54 |
|  | 6 | 24.1 | 7.51 |
|  | 7 | 24.0 | 7.50 |
|  | 8 | 24.0 | 7.46 |
|  | 9 | 23.9 | 7.45 |
|  | 10 | 23.9 | 7.44 |
|  | 11 | 23.8 | 7.42 |
|  | 12 | 23.8 | 7.39 |
| 2016 | 1 | 24.1 | 7.49 |
|  | 2 | 24.1 | 7.49 |
|  | 3 | 24.1 | 7.48 |
|  | 4 | 24.1 | 7.47 |
|  | 5 | 23.9 | 7.42 |
|  | 6 | 24.0 | 7.42 |
|  | 7 | 23.9 | 7.41 |
|  | 8 | 23.9 | 7.38 |
|  | 9 | 23.8 | 7.36 |
|  | 10 | 23.7 | 7.33 |
|  | 11 | 23.6 | 7.30 |
|  | 12 | 23.5 | 7.27 |
| 2017 | 1 | 19.8 | 6.12 |
|  | 2 | 19.8 | 6.11 |
|  | 3 | 19.8 | 6.10 |
|  | 4 | 19.7 | 6.08 |
|  | 5 | 19.7 | 6.06 |
|  | 6 | 19.6 | 6.05 |
|  | 7 | 19.6 | 6.02 |
|  | 8 | 19.5 | 6.01 |
|  | 9 | 19.5 | 5.99 |
|  | 10 | 19.5 | 5.98 |
|  | 11 | 19.2 | 5.91 |
|  | 12 | 19.2 | 5.89 |
| 2018 | 1 | 21.1 | 6.48 |
|  | 2 | 21.1 | 6.48 |
|  | 3 | 21.1 | 6.46 |
|  | 4 | 21.0 | 6.45 |
|  | 5 | 21.0 | 6.43 |
|  | 6 | 20.9 | 6.41 |
|  | 7 | 20.9 | 6.39 |
|  | 8 | 20.9 | 6.38 |
|  | 9 | 20.8 | 6.37 |
|  | 10 | 20.7 | 6.34 |
|  | 11 | 20.7 | 6.33 |
|  | 12 | 20.6 | 6.30 |

